# Determinants of antenatal care service utilization among adolescent mothers of (10-19) years of age at Kanyama level one hospital in Lusaka district

**DOI:** 10.1101/2024.11.05.24316786

**Authors:** Kenny Saka, Alice Ngoma Hazemba, Phoebe Bwembya

## Abstract

**Introduction:** The provision of adequate antenatal care services is crucial for promoting maternal and child health. Therefore, it is necessary to understand the factors influencing the use of these services, especially among special groups such as adolescents. Adolescents, defined as individuals between the ages of 10 and 19 years, face specific challenges and vulnerabilities during pregnancy and childbirth. Hence, it is vital to develop targeted interventions to improve their reproductive health outcomes. This study analyzed the factors associated with antenatal care utilization among adolescent mothers in Zambia.

**Methodology:** The study used a cross sectional study design. Data was collected using a structured questionnaire from adolescents who were 8 months pregnant or more and adolescents who were already mothers at Kanyama level one hospital in Lusaka. Descriptive statistics of individual characteristics were developed to describe the data. Associations between ANC utilization and independent variables was tested using the Chi-Square test and multivariate logistic regression.

**Results:** The proportion of antenatal care utilization in this study was 39.37%. At multivariate analysis, factors that were significantly associated with antenatal care utilization were past pregnant experience (OR = 0.35; 95% CI: 0.14 to 0.85) and partners education (OR = 1.38; 95% CI: 1.02 to 1.87). However, Partner education did not have a statistical significance between levels of education and ANC utilization. This could have been due to a small sample size. Other studies however, reviewed a significant statistical relationship between levels of education and adolescent ANC utilization.

**Conclusion:** The findings underscore a low proportion of antenatal care utilization among adolescent mothers. Particularly low proportion of ANC utilization was found in adolescents mothers who had never experienced pregnancy in the past.

## Introduction

Antenatal care (ANC) is a crucial component of maternal health services, designed to ensure the well-being of both mother and child during pregnancy. Among adolescent mothers aged 10-19 years, the utilization of these services is particularly critical due to the increased health risks associated with adolescent pregnancies^1^.

Adolescent pregnancy is a global phenomenon^1^. Every year in the low and middle income countries, an estimated 21 million girls aged 15–19 years become pregnant. A further 1.5 per 1000 girls aged 10-14 experience child birth globally. In Central and Southern Africa, adolescent birth rates are well above the global rate at 9 and 5 births per 1,000 adolescent girls aged 15-19 and 10-14^18^. In Zambia, 29% of adolescents aged 15 to 19 have begun childbearing, 24% have given birth while 5% are pregnant with their first child^2^.

Adolescent pregnancy have remained a major contributor to maternal and child mortality. Complications relating to pregnancy and childbirth are the leading cause of death for girls aged 10-19 globally^2^. Adolescents are also at significantly increased risk of delivering premature and low-birth-weight offspring. The risk of neonatal and perinatal mortality is even higher among adolescents less than 15 years. It is therefore, essential to protect the health of adolescent girls and their unborn children through ANC^3^.

However, the uptake of antenatal care visits among adolescent mothers in Zambia is low^4^. The 2021 Annual Health Statistical Report highlight that the national contribution of adolescents to the total antenatal attendances stood at 37.9%^5^. Hence, it is important to understand determinants of ANC to develop a targeted interventions to improve maternal and neonatal outcomes within this vulnerable population.

## Methods and Materials

### Study design

This was a cross sectional study. According to Thompson, cross sectional studies are one-time survey or observation of one or more groups of subjects. This is similar to taking a snap shot of these subjects at a single point in time. They are ideal for calculating simple prevalence rates. Investigators can also use this design to attempt to examine the natural history of phenomenon by performing cross-sectional observations^6^. Therefore, this study design was ideal for this study.

### Study population

The study population were adolescent girls who were 8 or more months pregnant and adolescent who were already mothers.

### Inclusion criteria

The study included adolescent who were already mothers and adolescents who were 8 or more months pregnant. The adolescents considered were only those found within the health facility premises.

### Exclusion criteria

Adolescents that were admitted at the health facility, referred to the general hospital and those that did not consent to participate in the study were excluded.

### Sampling Procedure

A simple random sampling method was used to collect data from participants. The study sample size was 475.

### Ethical approval

The study was approved by the University of Zambia Biomedical Research Ethics Committee (REF. No. 3624-2023). Permission for data collection was obtained from the Lusaka district medical office. Further, consent and assent forms were obtained from both the parents and the study participants.

### Data collection

Data were collected using a structured questionnaire on one on one interviews. A criterion of data completeness, timeliness, precision, integrity and validity was used and maximized to ensure data quality.

### Data Analysis

STATA version 14.2 was used for data analysis. Frequencies and percentages were generated and used to present categorical variables. To determine the association between ANC utilization and each independent variable, chi squared test and fishers exact test were used. This was dependent on the chi square assumptions being met. Logistic regression model was used to check for the relationship between the dependent and the independent variables. At multivariate regression analysis, an investigator led best fit model selection approach was used to select the best predictors of ANC in the multiple logistic regression model. Selection of the predictor variables was done by checking p-values. Variables found with larger p-value and not statistically significant were removed until statistically significant variables were left in the model.

## Results

### Individual, Structural and Health facility characteristics of adolescents at Kanyama level one hospital in Lusaka

Table 1 summarizes the results of the sample (n = 475). The sample comprised of adolescent mothers in the age range 10-19. The majority of the adolescent were in the age range 15-19 representing 97.89%. Less than a quarter (12.42%) reported they were married and 75.79% of the adolescents reported they were residing with their parents or guardians.

**Table 1:**
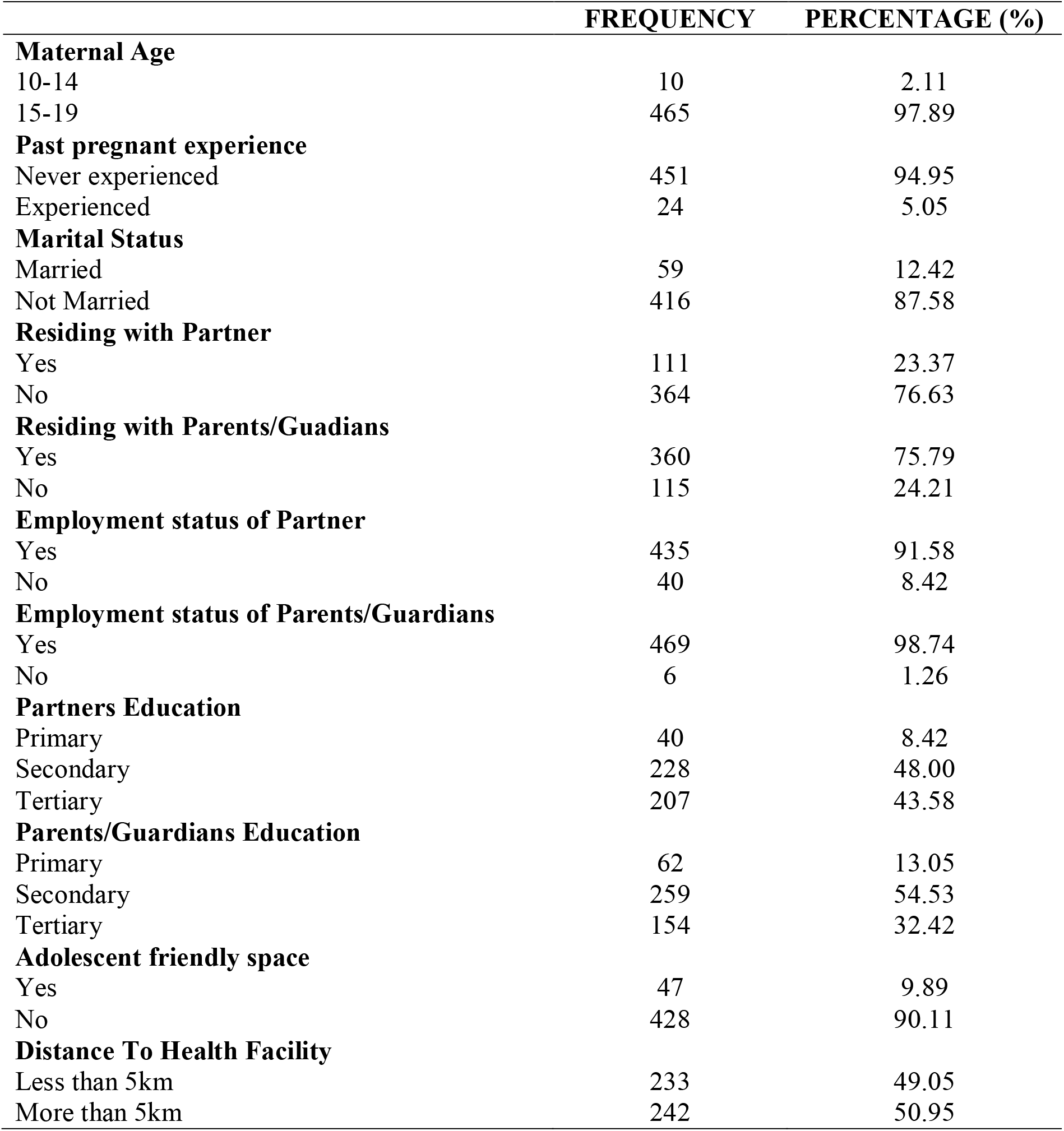
Individual, Structural and Health facility characteristics.

|  | FREQUENCY | PERCENTAGE (%) |
| --- | --- | --- |
| **Maternal Age** |  |  |
| 10-14 | 10 | 2.11 |
| 15-19 | 465 | 97.89 |
| **Past pregnant experience** |  |  |
| Never experienced | 451 | 94.95 |
| Experienced | 24 | 5.05 |
| **Marital Status** |  |  |
| Married | 59 | 12.42 |
| Not Married | 416 | 87.58 |
| **Residing with Partner** |  |  |
| Yes | 111 | 23.37 |
| No | 364 | 76.63 |
| **Residing with Parents/Guadians** |  |  |
| Yes | 360 | 75.79 |
| No | 115 | 24.21 |
| **Employment status of Partner** |  |  |
| Yes | 435 | 91.58 |
| No | 40 | 8.42 |
| **Employment status of Parents/Guardians** |  |  |
| Yes | 469 | 98.74 |
| No | 6 | 1.26 |
| **Partners Education** |  |  |
| Primary | 40 | 8.42 |
| Secondary | 228 | 48.00 |
| Tertiary | 207 | 43.58 |
| **Parents/Guardians Education** |  |  |
| Primary | 62 | 13.05 |
| Secondary | 259 | 54.53 |
| Tertiary | 154 | 32.42 |
| **Adolescent friendly space** |  |  |
| Yes | 47 | 9.89 |
| No | 428 | 90.11 |
| **Distance To Health Facility** |  |  |
| Less than 5km | 233 | 49.05 |
| More than 5km | 242 | 50.95 |

### ANC utilization among adolescent mothers

Table 2 shows the proportion of adolescent mothers who had utilized and who had not utilized ANC services. Utilized in this study is defined as having 4 or more contact visits to ANC services and not utilized is having 3 or less visits to ANC services among adolescent mothers who had 8 or more months pregnant or were already mothers. Only 39.37% of the adolescent mothers had utilized the ANC services.

**Table 2:** ANC utilization among adolescent mothers.

|  | **Pregnant Adolescents (n=475)** | **Percentage (%)** |
| --- | --- | --- |
| **ANC Utilization Proportion** |  |  |
| **Utilized** | 187 | 39.37 |
| **Not Utilized** | 288 | 60.63 |

### Factors associated with ANC utilization

Table 3 shows the chi square test of association between individual, structural, health facility factors and antenatal care utilization. There was an association between Past pregnant experience and antenatal care utilization at 95% CI, (0.005). There was also an association between Partners education and antenatal care utilization at 95% CI, (0.008). The rest of the variables were not statistically significant.

**Table 3:**
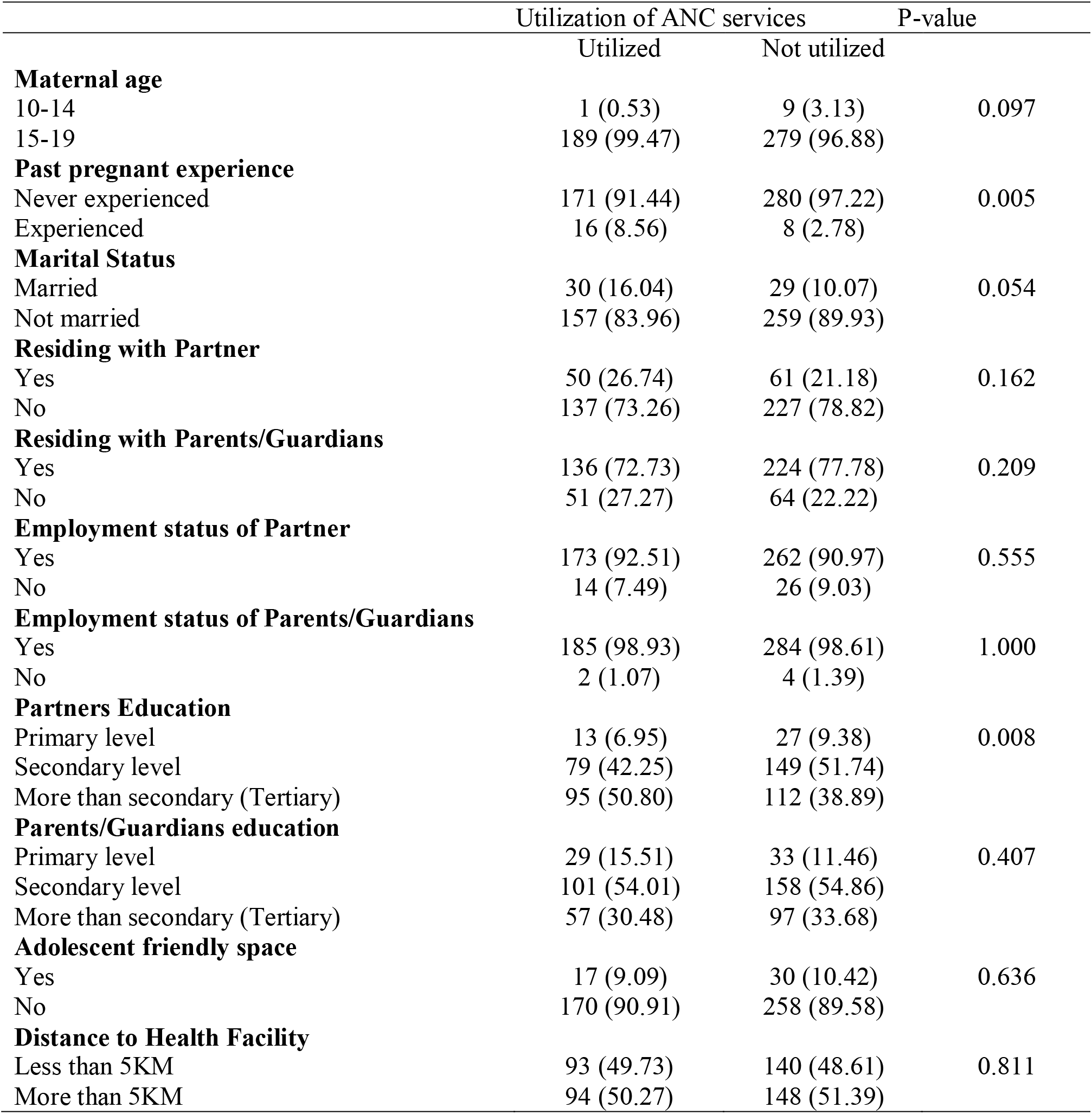
Factors associated with ANC utilization.

### Bivariate logistic regression analysis

The findings in table 4 shows that adolescents’ mothers who had never experienced past pregnant had lower odds of utilizing the ANC services. Their odds were 0.31 less likely to utilize ANC services compared to adolescents’ mothers who had experienced past pregnant (OR = 0.31; 95% CI: 0.13 to 0.73).

**Table 4:** Predictors of ANC utilization among adolescent mothers.

| Characteristics/factors | Study sample (%) | Proportional Odds Ratio (95%CI) | P-value |
| --- | --- | --- | --- |
| **Maternal age** |  |  |  |
| 10-14 | 2.11 | 1.0 |  |
| 15-19 | 97.89 | 5.99 (0.75-47.75) | 0.090 |
| **Past pregnancy experience** |  |  |  |
| Experienced | 5.05 | 1.0 |  |
| Never experienced | 94.95 | 0.31 (0.13-0.73) | 0.008 |
| **Marital Status** |  |  |  |
| Married | 12.42 | 1.0 |  |
| Not Married | 87.58 | 0.59 (0.34-1.01) | 0.056 |
| **Residing With Partner** |  |  |  |
| No | 76.63 | 1.0 |  |
| Yes | 23.37 | 1.36 (0.88-2.09) | 0.163 |
| **Residing with Parents/Guardians** |  |  |  |
| No | 24.21 | 1.0 |  |
| Yes | 75.79 | 0.76 (0.49-1.17) | 0.210 |
| **Employment status of Partner** |  |  |  |
| Not Employed | 8.42 | 1.0 |  |
| Employed | 91.58 | 1.23 (0.62-2.41) | 0.555 |
| **Employment status Parent/Guardians** |  |  |  |
| Not Employed | 1.26 | 1.0 |  |
| Employed | 98.74 | 1.30 (0.24-7.18) | 0.761 |
| **Partners education** |  |  |  |
| Primary | 8.42 | 1.0 |  |
| Secondary | 48.00 | 1.10 (0.54-2.25) | 0.792 |
| Tertiary | 43.58 | 1.76 (0.86-3.60) | 0.121 |
| **Parents/Guardians education** |  |  |  |
| Primary | 13.05 | 1.0 |  |
| Secondary | 54.53 | 0.73 (0.42-1.27) | 0.264 |
| Tertiary | 32.42 | 0.67 (0.37-1.21) | 0.186 |
| **Adolescent friendly space** |  |  |  |
| No | 90.11 | 1.0 |  |
| Yes | 9.89 | 0.86 (0.46-1.61) | 0.637 |
| **Distance To Health Facility** |  |  |  |
| Less than 5KM | 49.05 | 1.0 |  |
| More than 5KM | 50.95 | 0.96 (0.66-1.38) | 0.811 |

**Table 5:**
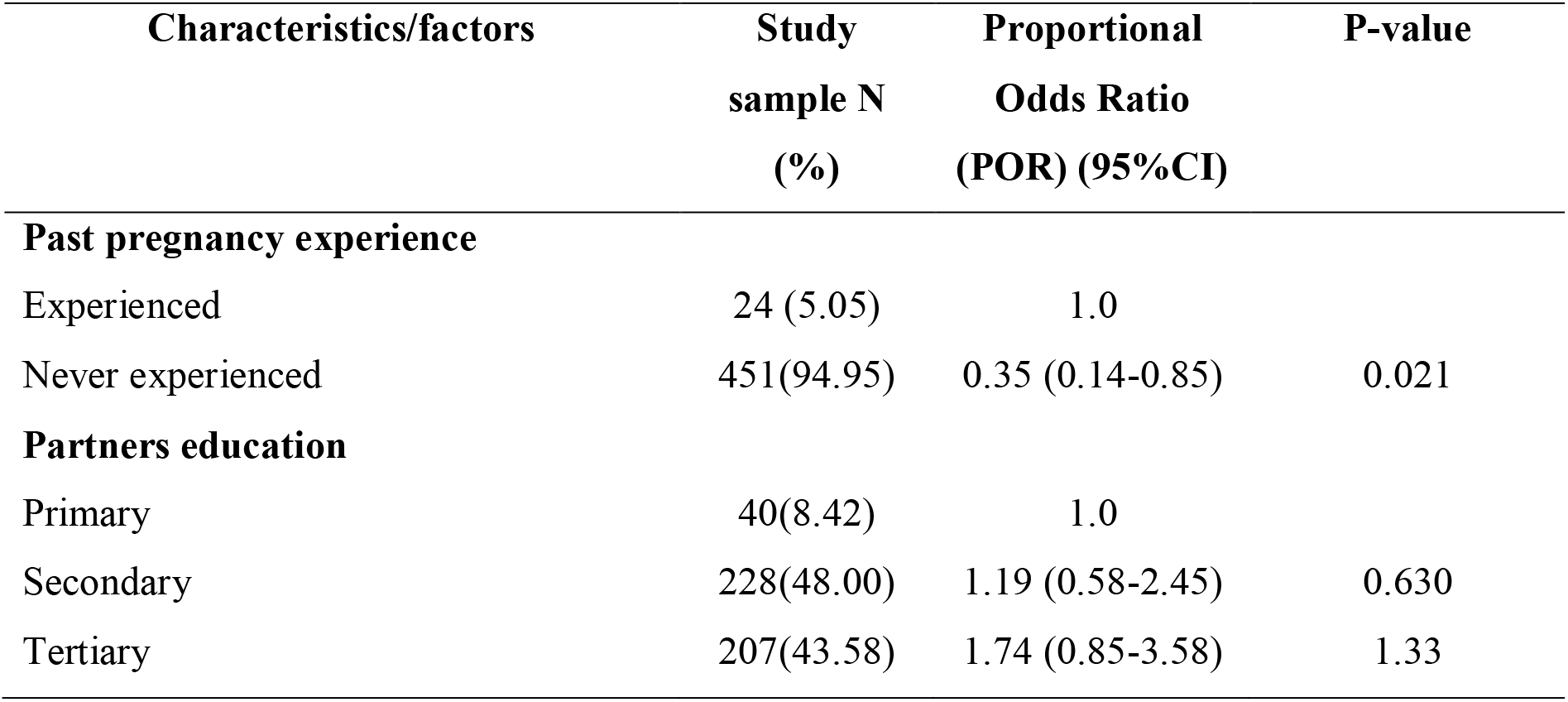
Multivariate analysis on factors associated with ANC utilization.

### Factors associated with ANC utilization

In this study, antenatal care utilization is associated with a number of individual, structural and health facility factors. However, in order to assess the contribution of all these factors to the overall variance, it was important at this stage to control for confounding by conducting a multiple logistic regression analysis

After adjusting for the effect of other factors, adolescents who had never experienced past pregnancy were less likely to utilize ANC services than adolescents who had experienced pregnancy before. Their odds were 0.35 less likely to utilize ANC services (OR = 0.35; 95% CI: 0.14 to 0.85).

## Discussion

Although antenatal care utilization is considered a success in Zambia, a notably low proportion of adolescent mothers sought antenatal care at Kanyama Level One Hospital in Lusaka. This figure reflects an ongoing challenge in achieving effective antenatal care utilization among adolescents. This finding aligns with the Annual Health Statistical Report (2022), which indicates that the contribution of adolescents to first antenatal attendances varies across provinces. Luapula had the highest attendance rate, while Lusaka had the lowest. Additionally, only four out of ten provinces, excluding six, recorded a significant contribution of adolescent mothers to first antenatal attendances. Based on these figures, the national picture suggests a concerning trend in antenatal care utilization among adolescent mothers across the provinces^7^.

There are variety of reasons that lead to adolescent mothers absconding ANC services. According to Kayemba (2023), attributed the absent from ANC services due to fear of pregnancy disclosure, long distance to health facility, lack of knowledge of pregnancy and ANC, travel costs to health facility and preference for traditional care to ANC^8^. Another study by Rahman et al, (2022), sites care seeking behavior during pregnancy. This is complex and is shaped by multiple factors of availability, accessibility, and acceptability of ANC services. Perceived usefulness and quality of ANC services significantly determine adolescent’s determination to return for subsequent visits. The study further sites that ANC services is more strongly associated with country’s GDP per capita than with coverage. Antenatal care (ANC) utilization varies significantly between low-income and upper-middle-income countries. Among adolescents, the primary reasons for lower ANC utilization include reduced bargaining power, lower expectations of care, and poorer health literacy^9^.

The current study found that adolescent mothers who had never experienced pregnancy before were less likely to utilize antenatal care services compared to those who had previous pregnancy experience. This is consistent with earlier research, which showed that adolescent mothers who had never been pregnant before had a lower prevalence of ANC utilization^10^. Another study by Skep found a significant association between past pregnancy experience and the use of a minimum of four ANC visits among adolescent mothers. It found that past pregnancy experience was positively associated with ANC use, with adolescent mothers who had experienced a previous birth being more likely to have at least four ANC visits than those whose most recent birth was their first^11^.

Chaibva reported that adolescent mothers who had never experienced pregnancy before had a lower prevalence of ANC utilization. Among adolescents that were interviewed, some feared disclosing their pregnancies to their parents or schools and preferred their pregnancies to be discovered when they were in labor. Some adolescents disclosed that they felt well, while others lacked money to pay ANC fees. Other reported reasons included limited knowledge about the benefits of ANC, reliance on traditional birth attendants, lack of required documents such as Zimbabwe national identity cards to register for ANC, and religious factors. This preference among pregnant adolescents could detrimentally influence their non-utilization of ANC services, with potentially hazardous effects for both mothers and babies^3^. Another study by Kayemba reported fear of pregnancy disclosure, long distances to health facilities, lack of knowledge about pregnancy and ANC, travel costs to health facilities, and a preference for traditional care over antenatal care services^4^.

Despite current study finding a significant association between partner’s education and antenatal care utilization, there was no significant association between levels of education and ANC utilization. This could have been attributed by a small sample size in this study. However, other studies such as that of Awingura found that adolescent mothers whose partners had secondary or more of education had a higher prevalence of ANC utilization compared to adolescent mothers whose partners had less than a secondary level of education. It further suggests that education positively impacts healthcare service utilization and increment of knowledge about specific issues hence recommends empowering adolescent mothers through education and decision-making to increase the utilization of maternal healthcare service^12^.

A study conducted by Christiana (2021) found that adolescent mothers with partners had some form of education were more likely to fully utilize ANC services compared to those whose partners had no education. This was attributed to the fact that education attainment is associated to higher awareness of the importance of ANC. No education attainment was seen to be associated with lower awareness of the importance of ANC^13^. Another study by Aduragbemi found that partner’s education was significant predictor for ANC utilization among adolescent mothers in Kenya. In this study, it was entirely conceivable that education was a significant factor for ANC utilization. However, it only suggested that education of partner enhances both resources and agency for ANC utilization in the interest of adolescent mothers as the two primary components of empowerment^14^.

Anaba 2022 noticed that partner’s education enabled mothers to have more access to health information, appreciate the causes of adverse pregnancy outcomes and the importance of ANC to the wellbeing of the mother and the unborn baby. In addition, partner’s education enabled adolescent mother to have greater autonomy to make decisions and financial access to quality healthcare^15^.

## Conclusion

The findings highlight a low rate of antenatal care (ANC) utilization among adolescent mothers, with especially low usage among those experiencing their first pregnancy. Various factors, such as lack of funds to cover ANC fees and limited knowledge about the benefits of ANC, may contribute to this low utilization. Therefore, it is crucial to implement strategies like making health facilities more accessible to adolescent mothers by addressing geographical and financial barriers and conducting community awareness campaigns to inform about the benefits of ANC and its importance for a healthy pregnancy.

## Data Availability

All data produced in the present study are available upon reasonable request to the authors

## Author’s contribution

The author was responsible for the Conceptualization, Methodology, Analyzing the data, interpretation of the findings and drafting the manuscript.

## Acknowledgement

The author wish to send sincere appreciation and acknowledge the invaluable contribution made by the lecturers at the University of Zambia, school of public health in particular DR Alice Ngoma Hazemba for supervising the work, Lusaka district medical office and Kanyama level one hospital for granting the permission to collect data and UNZABREC who granted ethical approval.

## Ethical approval

The study was approved by the University of Zambia Biomedical Research Ethics Committee (REF. No. 3624-2023). Prior to data collection, written permission was obtained from the Lusaka

District Medical Office. Additionally, consent and assent forms were provided to parents or next of kin, as well as to all study participants, to obtain their consent.

## Footnotes

1 ibid

2 1

3 10

4 8

